# Contemporary Endovascular Techniques for Cerebral Aneurysms: Germany-wide In-hospital Outcomes vs. Coiling (2013–2022)

**DOI:** 10.1101/2025.09.23.25336516

**Authors:** Konstantinos Vagkopoulos, Christian Haverkamp, Klaus Kaier, Jonas Werner, Mukesch Shah, Constantin von zur Mühlen, Jürgen Beck, Horst Urbach, Stephan Meckel

**Affiliations:** Department of Neuroradiology, Faculty of Medicine and Medical Center, University of Freiburg, Breisacher Straße 64, 79106 Freiburg, Germany; Institute of Digitalization in Medicine, Faculty of Medicine and Medical Center, University of Freiburg, Breisacher Straße 153, 79110 Freiburg, Germany; Institute of Medical Biometry and Statistics, Department of Methods in Clinical Epidemiology, Faculty of Medicine and Medical Center, University of Freiburg, Stefan-Meier-Straße 26, 79104 Freiburg, Germany; Department of Neurosurgery, Faculty of Medicine and Medical Center, University of Freiburg, Breisacher Straße 64, 79106 Freiburg, Germany; Departments of Cardiology and Angiology I, University Heart Center Freiburg, Faculty of Medicine and Medical Center, University of Freiburg, Breisacher Straße 64, 79106 Freiburg, Germany; Institute of Diagnostic and Interventional Neuroradiology, RKH Kliniken Ludwigsburg, Posilipostr. 4, 71640 Ludwigsburg, Germany

**Keywords:** intracranial aneurysm, nationwide analysis, clipping, coiling, balloon assisted coiling, stent assisted coiling, flow diversion, intrasaccular device

## Abstract

**Background:** Clinical and administrative studies suggest better functional outcomes after endovascular treatment (EVT) of intracranial aneurysms (IAs) compared to neurosurgical clipping (NSC). However, it remains unclear whether this applies to modern EVT techniques, such as balloon-assisted coiling (BAC), stent-assisted coiling (SAC), flow diversion (FD), or intrasaccular flow disruption (IFD). This study compares nationwide in-hospital outcomes of modern EVT methods and NSC with standard coiling (SC).

**Methods:** Administrative data from all German hospitals (2013–2022) were analyzed using billing codes for SC, BAC, SAC, FD, IFD, and NSC in ruptured and unruptured IAs. Primary outcomes included functional independence (discharge type), poor outcomes (US Nationwide Inpatient Sample-Subarachnoid Hemorrhage Outcome Measure [NIS-SOM]), and in-hospital mortality. Propensity score weighting was used for comparisons.

**Results:** A total of 77,684 procedures were analyzed (46.8% ruptured, 53.2% unruptured). In ruptured IAs, SAC, FD, and NSC were associated with lower functional independence (p=0.001, p=0.007, p<0.001) and higher mortality (p<0.001, p=0.001, p=0.032). Poor outcomes were more frequent after SAC (p=0.001) and NSC (p<0.001). In unruptured IAs, functional independence improved with BAC (p=0.036), SAC (p=0.045), and IFD (p<0.001), but decreased with NSC (p=0.017). Poor outcomes were less frequent with IFD (p<0.001), and mortality was lower with NSC (p=0.020) and IFD (p=0.003).

**Conclusions:** Nationwide data from Germany reveal significant differences between EVT techniques and NSC for IA treatment. In ruptured IAs, SAC, FD, and NSC were associated with worse outcomes compared to SC. In unruptured IAs, BAC, SAC, and IFD improved functional outcomes, while NSC was linked to decreased functional outcomes. Notably, IFD consistently demonstrated superior functional outcomes despite limited utilization. Given the limitations of billing data, these findings suggest a potential shift favoring IFD as a safer treatment option in unruptured IAs.

## Introduction

Intracranial aneurysms (IAs) as acquired vascular lesions are a significant health concern, with the potential to cause devastating neurological damage or death if left untreated and rupture.^1^ Since the first clip-occlusion of an IA by Walter E. Dandy in 1937,^2^ neurosurgical clipping (NSC) has been widely used and evolved by introduction of microscopic surgery. Although further advances in operative techniques and approaches have occurred, NSC as an invasive procedure is often associated with intraoperative and postoperative complications.^3^ Endovascular treatment (EVT) of IAs which has emerged in the 1990s with the advent of the Guglielmi detachable coiling (GDC), established a new era of neurointervention, with favorable clinical results of standard coiling (SC) compared to NSC for ruptured IAs associated with lower risk of mortality and morbidity.^4^ ^5^ Although SC was proven to be an effective treatment in many IA cases, high rates of aneurysm recanalization and re-treatments were subsequently observed.^6^

Since then, EVT technology for aneurysms has rapidly emerged with many new devices and techniques being developed during the past two decades. Their goal was to improve ease and safety of EVT as well as to prevent aneurysm regrowth particularly focusing on wide-necked or other complex aneurysm morphologies being notoriously difficult to treat and highly prone to recur after SC. These techniques include balloon-assisted coiling (BAC),^7^ ^8^ stent-assisted coiling (SAC),^9^ flow-diversion (FD),^10^ ^11^ and several devices for intrasaccular flow disruption (IFD) such as the WEB device.^12^ ^13^ Their safety and efficacy have been demonstrated in many retrospective as well as prospective cohort or registry studies.^14^ However, large- scale or randomized data comparing different EVT methods and NSC is still lacking.

Recently, we have reported the in-hospital outcomes of > 90 000 IA aneurysm treatments from a nationwide German administrative analysis between 2008-2019 demonstrating higher rates of functional independence and lower rates of poor outcomes at discharge with equal mortality for EVT in general compared to NSC.^15^

Herein, we aimed to study nationwide outcomes for all contemporary EVT techniques based on an administrative dataset of all German hospital inpatients treated between 2013-2022 for unruptured and ruptured IAs. Our analysis included in-hospital safety outcomes and mortality from treatments with BAC, SAC, FD, IFD, and clipping compared to SC as a reference.

## Methods

### Data source, standard protocol approvals, registrations and patient consents

Starting from 2005, the German Federal Statistical Office (DESTATIS) has been providing summarized and anonymous results concerning specific inquiries related to billing data in Germany’s hospitals. This information is based on the diagnosis related group system and encompasses diagnoses (classified under the International Classification of Diseases, Tenth Revision, ICD-10) as well as performed procedures (coded according to the Operation and Procedure Classification System [OPS]).

These data are accessible upon request and can be utilized by anyone, if they adhere to the specified terms of use. To this study, only these publicly available and aggregated routine health data were examined. Therefore, obtaining additional approval from a local ethics committee or patient consent was not necessary. The analysis of the data adheres to the guidelines outlined in the RECORD guideline.

### Study population, treatment modalities, and group allocations

By reviewing the period from 2013-2022 based on the same data source as recently described by Haverkamp et al., we identified inpatient cases treated for IA according to the case selection algorithm outlined in Figure 1. In a first step we identified the cases with a billing code (OPS code) for EVTs (SC, BAC, SAC, FD, IFD) or NSC and a diagnosis code (ICD-10) of subarachnoid hemorrhage (SAH), intracerebral hemorrhage (ICH), or intracranial aneurysm (IA) (see also flowchart in Figure 1, detailed description of utilized ICD-10 and OPS codes in Table S1). Thereby, multiple IA treatments consisting of separated hospital re-admissions within a single patient were regarded as separate inpatient cases.

**Figure 1:**
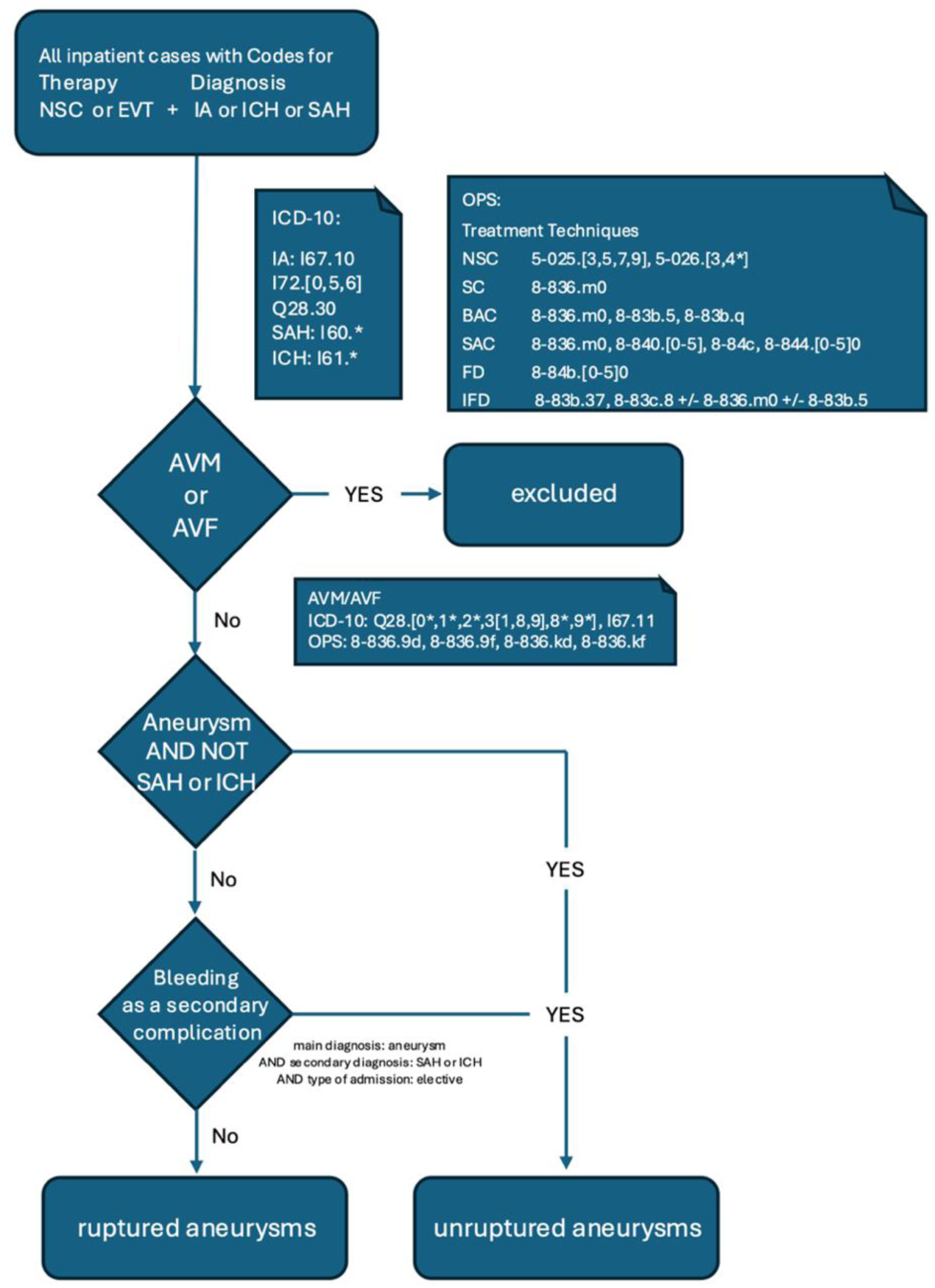
Flow chart for cohort definition of inpatient cases with treatment of ruptured and unruptured intracranial aneurysms. AVF: arteriovenous fistula, AVM: arteriovenous malformation, BAC: balloon-assisted coiling, EVT: endovascular treatment, FD: flow diverter, IA: intracranial aneurysm, ICH: intracranial aneurysm, IFD: intrasaccular flow disruption, NSC: neurosurgical clipping, SAC: stent-assisted coiling, SAH subarachnoid hemorrhage, SC: standard coiling

Then we excluded cases with additional diagnostic codes for other cerebrovascular conditions, such as arteriovenous malformations or fistulas, which may also lead to intracranial hemorrhage and may be treated using similar procedures, like coil embolization, to prevent misclassification.

The remaining cases were categorized into two groups: ruptured and unruptured IAs, based on ICD-10 principal and secondary diagnoses and the hospital admission type as defined in § 301 SGB V. The ruptured group included those with a coded diagnosis of SAH or ICH, while cases treated for IA without a hemorrhage diagnosis were assigned to the unruptured group.

To further reduce misclassification, cases with elective IA treatment and a secondary diagnosis of intracranial hemorrhage were considered procedural complications and assigned to the unruptured IA group. In the German DRG (diagnosis related group) system the primary diagnosis reflects the condition responsible for the hospitalization, hence periprocedural hemorrhage in elective cases should not be coded as the principal diagnosis.

Comorbidities were assessed by the Elixhauser Comorbidity Index (van Walraven’s weights).^16^

Like other nationwide inpatient samples our dataset was lacking traditional scores (Hunt and Hess, Fisher etc.) to evaluate the severity of aneurysmal subarachnoid hemorrhage (SAH). For this reason, derived from diagnostic and procedural codes we used the employed surrogate marker NIS-SSS (Nationwide Inpatient Sample- Subarachnoid Hemorrhage Severity Score akin to Hunt and Hess) as described by Washington et. al. (see Table S2).^17^ However, we did not use NIS-SSS as an outcome or parameter to adjust the outcomes of ruptured IA treatments since many of the diagnoses included in this score could be related to both sequelae of IA rupture with SAH/ICH as well as complications arising from IA treatment itself such as coma, aphasia, paraplegia etc.

### Outcomes

Functional independence at hospital discharge and in-hospital mortality were defined as the primary outcomes, initially determined by the type of hospital discharge (discharge types are summarized in Table S3). To measure functional independence as an ordinal measure, the discharge types were further divided into three categories of functional outcome at hospital discharge:

1. Good outcome (defined as discharge to home).
2. Intermediate outcome (indicating that patients required further therapy after discharge).
3. Poor outcome (intrahospital death).

Additionally, the study incorporated the Nationwide Inpatient Sample-Subarachnoid Hemorrhage Outcome Measure (NIS-SOM)^17^ as a measurement for clinical outcome (akin to modified Rankin Scale [mRS], see Table S4):

- Good, discharge to home/rehabilitation facility and/or other hospital

- Poor, for in-hospital mortality, discharge to a nursing facility, extended care facility/hospice, placement of a tracheostomy tube and/or gastrostomy tube.

Secondary outcomes comprised the duration of inpatient stay, duration of mechanical ventilation, rate of prolonged ventilation (more than 48 hours), and treatment costs based on the case mix revenues from German Diagnosis-Related Group (DRG) system.

### Statistical Evaluation

Since patients were not randomized towards the different treatment options, potential confounding factors were considered using the propensity score methods. Thereby, inverse probability weighting was applied. The propensity score is defined as the conditional probability of an individual for being in the treatment group, given a group of observed covariates. For the propensity score estimations, we fit multinomial logistic regression models with the treatment category SC as the reference category and controlling for 6 predetermined covariates (age, sex, Elixhauser Comorbidity index, atrial fibrillation, hyperlipidemia, and history of stroke), and the treatment year as categorical covariate. For the outcome models, we chose poisson regression models for the inverse probability weighting approach. Thereby, cluster-robust standard errors were used to account for the correlation of error terms of patients treated in the same hospital. Exponentiated coefficients of these weighted poisson regression models may be interpreted as relative risks in case of dichotomous endpoints or incidence rate ratios in case of continuous endpoints (ordinal outcome). For length of hospital stay and reimbursement, weighted linear regression models were applied. All regression models were specified using cluster robust standard errors to account for the clustering of patients within the same hospitals.

Standardized mean difference (SMD) was used as a statistic to examine the balance of covariate distribution between treatment groups after inverse probability weighting. A SMD<0.1 is conserved appropriate.

## Results

77 684 inpatient cases with IA treated in all German hospitals between 2013 and 2022 were identified (46.8% were classified as unruptured and 53.2% as ruptured). The distribution of treatment techniques for all cases was as follows: SC 28.3%, BAC 8.6%, SAC 11.4%, FD 13.0%, IFD 1.8%, and NSC 31.1%. Cases with combined use of EVT and NSC (1.8%) within the same hospital stay were excluded from further statistical analysis.

The overall portion of female cases was 65.7% and 72.1% in the ruptured IA and unruptured IA group, respectively. The mean age was 56 years (56.4 years in the ruptured IA group of, and 56.6 years in the unruptured IA group). Baseline characteristics for all treatment groups are summarized in Table 1.

**Table 1:** Baseline characteristics.

| **Ruptured aneurysms** |  |  |  |  |  |  |
| --- | --- | --- | --- | --- | --- | --- |
|  | SC | BAC | SAC | FD | IFD | NSC |
| N<br>(% of main groups) | 13 685<br>(38.8%) | 3 806<br>(10.8%) | 1 948<br>(5.5%) | 1 392<br>(4.0%) | 1 459<br>(4.1%) | 12 924<br>(36.7%) |
| Age<br>(years, mean ± SD) | 56.75<br>±13.72 | 55.81<br>±13.47 | 57.9<br>±13.3 | 56.76<br>±13.27 | 56.92<br>±13.16 | 55.80<br>±13.54 |
| Sex: Female | 66.7% | 68.4% | 66.3% | 61.5% | 65.5% | 65.7% |
| Elixhauser<br>comorbidity index<br>(mean ± SD) | 5.92<br>±5.13 | 6.09<br>±5.37 | 6.44<br>±5.14 | 6.08<br>±5.4 | 5.63<br>±5.14 | 6.73<br>±5.34 |
| Atrial Fibrillation | 7.0% | 6.8% | 7.8% | 8.3% | 6.0% | 6.3% |
| Hyperlipidemia | 7.6% | 9.2% | 8.7% | 8.1% | 6.0% | 7.3% |
| Stroke history | 0.7% | 0.9% | 1.2% | 1.2% | 0.7% | 0.6% |
| NIS-SSS<br>(mean ± SD) | 14.41<br>±21.7 | 17<br>±26.96 | 15.90<br>±23.69 | 15.7<br>±24.01 | 12.98<br>±20.22 | 15.36<br>±20.94 |
| NIS-SSS>7 | 62.5% | 58.7% | 66.1% | 63.1% | 59.8% | 70.4% |
| **Unruptured aneurysms** |  |  |  |  |  |  |
|  | SC | BAC | SAC | FD | IFD | NSC |
| N<br>(% of main groups) | 8 360<br>(20.3%) | 2 840<br>(6.9%) | 6 933<br>(16.9%) | 8 690<br>(21.1%) | 3 030<br>(7.4%) | 11 264<br>(27.4%) |
| Age<br>(years, mean ± SD) | 56.75<br>±12.13 | 55.29<br>±12.61 | 57.00<br>±11.55 | 56.68<br>±12.63 | 58.48<br>±11.26 | 55.86<br>±11.19 |
| Sex: Female | 70.8% | 73.3% | 72.0% | 75.0% | 69.2% | 72.5% |
| Elixhauser<br>comorbidity index<br>(mean ± SD) | 1.62<br>$\pm 3.67$ | 1.60<br>$\pm 3.59$ | 1.39<br>$\pm 3.41$ | 1.44<br>$\pm 3.47$ | 1.40<br>$\pm 3.49$ | 2.76<br>$\pm 4.54$ |
| Atrial Fibrillation | 4.0% | 4.0% | 3.0% | 3.4% | 4.2% | 3.7% |
| Hyperlipidemia | 14.1% | 11.8% | 11.5% | 12.5% | 11.2% | 12.5% |
| Stroke history | 2.5% | 3.0% | 2.9% | 2.3% | 2.4% | 2.2% |
| NIS-SSS<br>(mean ± SD) | N/A | N/A | N/A | N/A | N/A | N/A |
| NIS-SSS>7 | N/A | N/A | N/A | N/A | N/A | N/A |

SC: standard coiling, BAC: balloon assisted coiling, SAC: stent-assisted coiling, FD: flow diverter, IFD: intrasaccular flow diversion, NSC: neurosurgical clipping, N: Number, NIS-SSS: Nationwide Inpatient Sample-Subarachnoid hemorrhage Severity Score, SD: standard deviation

In ruptured IAs group, the distribution of treatment methods was as follows: SC 37.6% (13 658), BAC 10.5% (3 806), SAC 5.4% (1 948), FD 3.8% (1 392), IFD 4.0% (1 459), and NSC 35.6% (12 924 cases) respectively. A combination of NSC and EVT within the same inpatient case was performed in 3.2% (1 155 cases).

Whereas unruptured IA cases were treated with SC 20.2% (8 360), BAC 6.9% (2 840), SAC 16.8% (6 933), FD 21.0% (8 690), IFD 7.3% (3 030) and NSC 27.2% (11 264), respectively. Here, a combined endovascular and neurosurgical treatment within the same inpatient case was performed in 0.5% (225 cases).

### Outcomes in Ruptured IA Group

All primary and secondary outcomes after treatments performed for ruptured IAs are summarized in Table 2. In unadjusted analysis (prior to propensity score weighting), all primary outcomes differed significantly among all analyzed treatment techniques (p<0.001): Functional independence ranged between 30.1% (NSC) and 40.2% (BAC); poor clinical outcome (NIS-SOM) varied between 37.2% (BAC) and 49.8% (NSC) and in-hospital mortality varied between 16.2% (IFD) and 24.5% (SAC).

**Table 2:** Outcomes of Different Treatment Methods in Ruptured Aneurysms.

|  | SC | BAC | SAC | FD | IFD | NSC |
| --- | --- | --- | --- | --- | --- | --- |
| N | 13 658<br>(38.8%) | 3 806<br>(10.8%) | 1 948<br>(5.5%) | 1 392<br>(4.0%) | 1 459<br>(4.1%) | 12 924<br>(36.7%) |
| **Primary outcomes** |  |  |  |  |  |  |
| Good (discharge to home) | 37.9% | 40.2% | 34.7% | 34.8% | 39.5% | 30.1% |
| Intermediate (further hospital/rehab/nursing home) | 44.1% | 43.2% | 40.9% | 42.3% | 44.3% | 50.8% |
| Poor (intrahospital death) | 18.0% | 16.6% | 24.5% | 22.9% | 16.2% | 19.1% |
| NIS-SOM (mRS>2-3) | 39.5% | 37.2% | 45.2% | 44.3% | 37.7% | 49.8% |
| **Secondary outcomes** |  |  |  |  |  |  |
| Length of stay (days, mean ± SD) | 24.48<br>±19.99 | 25.95<br>±22.25 | 24.12<br>±24.54 | 24.53<br>±21.83 | 22.52<br>±15.06 | 28.25<br>±26.69 |
| Ventilation (hours, mean ± SD) | 195.47<br>±275.87 | 193.80<br>±287.95 | 212.87<br>±292.03 | 215.98<br>±309.63 | 163.92<br>±239.85 | 259.98<br>±297.73 |
| Ventilation >48h | 50.1% | 47.6% | 54.7% | 54.5% | 45.0% | 63.0% |
| Costs (Euro, mean ± SD) | 34<br>787.21<br>±30<br>561.12 | 35<br>859.80<br>±33<br>624.92 | 37<br>731.61<br>±34<br>295.97 | 36<br>816.04<br>±31<br>406.69 | 31<br>861.91<br>±22<br>597.91 | 43<br>117.60<br>±38<br>271.29 |

SC: standard coiling, BAC: balloon-assisted coiling, SAC: stent-assisted coiling, FD: flow diverter, IFD: intrasaccular flow diversion, NSC: neurosurgical clipping, N: number, NIS-SOM: Nationwide Inpatient Sample-Subarachnoid hemorrhage Outcome Measure, SD: standard deviation

After propensity score weighting (using SC as reference, detailed analysis in Table S5), functional independence was found significantly less frequently after SAC (RR=1.097, 95%-CI 1.041 to 1.156, p=0.001), FD (RR=1.104, 95%-CI 1.028 to 1.186, p=0.007), and NSC (RR=1.109, 95%-CI 1.072 to 1.147, p<0.001). Similarly, the rates of poor outcomes were higher after SAC (RR=1.115, 95%-CI 1.049 to 1.186, p=0.001) and NSC (RR=1.241, 95%-CI 1.176 to 1.309, p<0.001), and in-hospital mortality was increased after SAC (RR=1.331, 95%-CI 1.211 to 1.462, p<0.001), FD (RR=1.294, 95%-CI 1.114 to 1.504, p=0.001), and NSC (RR=1.083, 95%-CI 1.007 to 1.164, p=0.032).

In contrast, no significant differences were observed in all primary outcomes between SC, BAC, and IFD.

Regarding secondary outcomes, IFD was associated with a statistically significant reduction in hospital stay (by 2 days, 95%-CI -3.811 to -0.233, p=0.027) and ventilation time (by 33.6 hours, 95%-CI -64.64 to -2.63, p=0.034; prolonged ventilation over 48 hours was less frequent RR 0.904, 95%-CI 0.818 to 1.000, p=0.049) compared to SC (propensity score weighted analysis). Conversely, NSC prolonged hospital stay (by 3 days, 95%-CI 1.660 to 4.366, p<0.001) and ventilation time (by 54.7 hours, 95%-CI 40.28 to 69.15, p<0.001; prolonged ventilation over 48 hours was more frequent RR 1.226, 95%-CI 1.176 to 1.279, p<0.001). SAC was associated with an extended rate of prolonged ventilation over 48 hours (RR 1.063, 95%-CI 1.008 to 1.122, p=0.025).

In terms of financial impact, SAC (+€2,139, 95%-CI 274 to 4003, p=0.025) and NSC (+€7,115, 95%-CI 5242 to 8987, p<0.001) were more expensive than SC, while IFD led to a cost reduction (-€3,200, 95%-CI -6276 to -125, p=0.041).

### Outcomes in unruptured intracranial aneurysms

Table 3 lists all primary and secondary outcomes following different treatment modalities in unruptured IAs. Differences were observed in unadjusted analysis between all six treatment techniques for the primary endpoints. The rates of functional independence ranged from 86.4% (NSC) to 95.5% (IFD), the rates of poor clinical outcome ranged from 1.1% (IFD) to 4.3% (NSC), and mortality varied between 0.4% (IFD) and 1.4% (SC).

**Table 3:** Outcomes of Different Treatment Methods in Unruptured Aneurysms.

|  | SC | BAC | SAC | FD | IFD | NSC |
| --- | --- | --- | --- | --- | --- | --- |
| N | 8 360<br>(20.3%) | 2 840<br>(6.9%) | 6 933<br>(16.9%) | 8 690<br>(21.1%) | 3 030<br>(7.4%) | 11 264<br>(27.4%) |
| **Primary outcomes** |  |  |  |  |  |  |
| Good (discharge to home) | 91.9% | 94.0% | 94.5% | 94.0% | 95.5% | 86.4% |
| Intermediate (further hospital/rehab/nursing home) | 6.7% | 4.7% | 4.5% | 4.7% | 4.1% | 12.3% |
| Poor (intrahospital death) | 1.4% | 1.3% | 1.0% | 1.3% | 0.4% | 1.3% |
| NIS-SOM (mRS>2-3) | 3.3% | 2.5% | 2.1% | 2.5% | 1.1% | 4.3% |
| **Secondary outcomes** |  |  |  |  |  |  |
| Length of stay (days, mean ± SD) | 7.08<br>$\pm 8.82$ | 6.67<br>$\pm 7.29$ | 6.15<br>$\pm 6.57$ | 6.25<br>$\pm 6.61$ | 5.58<br>$\pm 5.21$ | 12.55<br>$\pm 11.61$ |
| Ventilation (hours, mean ± SD) | 16.29<br>$\pm 97.90$ | 12.54<br>$\pm 75.26$ | 8.01<br>$\pm 63.09$ | 8.89<br>$\pm 74.88$ | 4.65<br>$\pm 43.05$ | 25.41<br>$\pm 126.65$ |
| Ventilation >48h | 4.3% | 3.6% | 2.2% | 2.6% | 1.5% | 6.6% |
| Costs (Euro, mean ± SD) | 11<br>011.19<br>$\pm 11$<br>870.58 | 10<br>659.57<br>$\pm 8$<br>883.54 | 14<br>645.21<br>$\pm 7$<br>981.45 | 10<br>824.28<br>$\pm 8$<br>228.90 | 10<br>188.70<br>$\pm 5$<br>493.19 | 15<br>354.40<br>$\pm 14$<br>693.78 |

SC: standard coiling, BAC: balloon-assisted coiling, SAC: stent-assisted coiling, FD: flow diverter, IFD: intrasaccular flow diversion, NSC: neurosurgical clipping, N: Number, NIS-SOM: Nationwide Inpatient Sample-Subarachnoid hemorrhage Outcome Measure, SD: standard deviation

After propensity weighting (with SC as a reference, detailed analysis in Table S6), functional independence was less often observed after NSC (RR=1.263, 95%-CI 1.042 to 1.531; p=0.017), and more often after BAC (RR=0.787, 95%CI 0.629 to 0.985; p=0.036), SAC (RR=0.743, 95%CI 0.556 to 0.994; p=0.045) and IFD (RR=0.546, 95%CI 0.408 to 0.73; p<0.001). Rates of poor clinical outcome were lower for IFD (RR=0.370, 95%CI 0.233 to 0.588; p<0.001). Mortality was lower after NSC (RR=0.726, 95%CI 0.554 to 0.951, p=0.02), and IFD (RR=0.374, 95%CI 0.171 to 0.704, p=0.003).

Regarding the secondary outcomes, length of hospital stay was shortened in cases with SAC (by 0.79 days, 95%CI -1.507 to -0.069, p=0.032) and IFD (by 1.76 days, 95%CI -1.987 to -0.342, p=0.006) and prolonged after NSC (by 4.6 days, 95%CI 3.828 to 5.391, p<0.001). Ventilation duration was reduced after SAC (by 7.3 hours, 95%CI -12.74 to -1.86, p=0.009) and after IFD (by 11 hours, 95%CI -17.26 to -4.93, p<0.001).

The rate of prolonged ventilation over 48 hours was reduced after SAC (RR 0.602, 95%-CI 0.428 to 0.848, p=0.004), FD (RR 0.690, 95%-CI 0.490 to 0.970, p=0.033) and IFD (RR 0.365, 95%-CI 0.252 to 0.529, p<0.001).

In terms of costs, SAC (+€3 659, 95%CI 2 971 to 4 348, p<0.001) and NSC (+€3 454, 95%CI 2 893 to 4 015, p>0.001) were more expensive.

## Discussion

This nationwide study presents a comprehensive real-world analysis of in-hospital outcomes associated with various endovascular treatment (EVT) modalities for intracranial aneurysms (IAs) in Germany between 2013 and 2022—a period marked by the widespread adoption of modern EVT techniques. It offers comparative insights into major EVT methods and neurosurgical clipping (NSC) against the benchmark of simple coiling (SC) in both ruptured and unruptured IAs.

SC and NSC were the most commonly employed treatment approaches, each accounting for approximately one-third of cases. Advanced modalities such as BAC, SAC, FD, and IFD were used less frequently, likely reflecting limited adoption outside high-volume centers, anatomical constraints, device-related considerations (e.g., need for dual antiplatelet therapy), need for training technical skills, and limited long-term data.

Historically, NSC has been the cornerstone of IA management, evolving with microsurgical advancements. However, SC has become the most widely adopted modality, supported by landmark studies like the International Subarachnoid Aneurysm Trial (ISAT)^4^, which showed a 23% relative risk reduction in death or disability at one year for SC versus NSC in ruptured IAs. Though ISAT had limitations—including selection bias and reduced generalizability—its findings were reinforced by trials such as the Barrow Ruptured Aneurysm Trial (BRAT)^5^, and recent meta-analyses (e.g., Delgado et al.)^18^, which reported higher independence and lower mortality with SC.

Our previous analysis (2007–2019) similarly demonstrated superior functional outcomes and comparable mortality for EVT versus NSC.^15^ The current analysis confirms these findings: for ruptured IAs, NSC was associated with lower functional independence, higher rates of poor outcomes, and increased mortality after propensity score adjustment. Though, lower functional outcomes at discharge after NSC may be in part related to the craniotomy procedure and are likely subject to further improvement after rehabilitation at long-term assessment.

Despite its efficacy, SC is challenged by durability concerns. Studies like MAPS have highlighted suboptimal complete occlusion rates (42% at one year) and substantial retreatment rates.^19^ These limitations, especially in anatomically complex aneurysms (e.g., wide-neck, fusiform, blister), have driven the development of adjunctive techniques.

### Balloon-Assisted Coiling

BAC offers temporary support for coil placement, particularly in wide-necked aneurysms. Data from ATENA (for unruptured IAs)^7^ and CLARITY (for ruptured IAs)^8^ have shown BAC achieves comparable or improved occlusion rates over SC, with no increase in morbidity or mortality. Our findings align with this: in ruptured IAs, BAC had no significant safety disadvantage compared to SC. In unruptured IAs, BAC showed higher functional independence but was underutilized—possibly due to concerns about long-term stability compared to SAC or FD.

### Stent-Assisted Coiling

SAC provides structural support for treating complex wide-necked aneurysms (e.g. by utilizing single or multiple stents in configurations such as half-T, T, or Y), especially in unruptured cases. While offering higher occlusion durability, SAC is associated with increased thromboembolic risks and requires dual antiplatelet therapy (DAPT).^20^ ^21^ ^22^ The latter raises concerns in ruptured aneurysms due to the risk of hemorrhagic complications during the acute SAH phase.^23^ In our cohort, SAC was used infrequently (5.4%) in ruptured cases and correlated with worse outcomes—including lower functional independence, increase in poor outcome, and higher ventilation times. Conversely, SAC in unruptured IAs was associated with similar or slightly improved outcomes compared to SC, with shorter hospitalization and reduced ventilation.

### Flow-Diversion

FD represents a transformative approach in EVT, especially for large, fusiform, or sidewall aneurysms. Since the first introduction of Pipeline embolization device (Covidien, Mansfield, MA, USA) being the first FD to be approved by the FDA in 2011^10^ ^11^, this approach was initially limited to select internal carotid artery cases. However, the introduction of new-generation FDs (e.g., Silk Vista Baby) and surface modifications^24^ ^25^ has broadened their use—including in distal or smaller vessels and under single antiplatelet therapy (SAPT). Moreover, FDs have shown excellent long-term efficacy with complete or near complete occlusion rates specifically for unruptured aneurysms between 77-96% (1-5 years outcomes).^26^ Nevertheless, FD carries risk: ischemic events, delayed rupture, and the burden of antiplatelet therapy in acute SAH remain serious concerns.^27^ Our findings reflect this—FD use in ruptured IAs was rare (3.8%) and associated with poorer outcomes and increased mortality. Thus, FDs should be reserved for ruptured IAs which are not suitable for treatment by other EVT or surgical techniques (e.g. blister or dissecting aneurysms).

In contrast, FD was used in over 20% of unruptured cases and showed equivalent in-hospital outcomes to SC, underlining its safety in carefully selected patients as an highly effective approach for various types of unruptured IAs with challenging morphologies (e.g. wide-necked, side-wall, distal, fusiform, giant, partially thrombosed, etc.).

### Intrasaccular Flow-Disruption

IFDs, particularly the Woven EndoBridge (WEB), offer an alternative treatment option for wide-necked bifurcation aneurysms by eliminating the need for stents and DAPT. GCP (Good Clinical Practice) trials have consistently shown low morbidity (1-3%) and zero mortality for unruptured IAs treated with WEB.^28^ ^12^ Moreover, WEB demonstrated no rebleeding and low rates of long-term morbidity and mortality for ruptured IAs in a European multicenter GCP study (CLARYS)^13^ and in a multicenter retrospective US study^29^. Our study likely reflects primarily WEB use, as newer IFDs like Contour^30^ ^31^ and Artisse^32^ were introduced late in the study period. Despite limited adoption during our analysis (4% ruptured, 7.3% unruptured), IFDs demonstrated high safety: lowest mortality and poor outcome rates, superior functional independence, shorter ventilation times, and reduced hospital stays.

These findings support the growing consensus that IFDs are a safe and effective option for appropriately selected wide-neck bifurcation aneurysms. The low rates of IFD utilization likely relate to their novelty that still precludes widespread use outside of high-volume centers, and to device-specific anatomical, technical and size limitations (e.g. not suitable for large/giant or fusiform aneurysms).

### Strengths and limitations

A key strength of this study is its use of the nationwide DESTATIS database, enabling a comprehensive assessment of real-world in-hospital outcomes across all levels of care in Germany over a 10-year period. This broad dataset enhances generalizability by capturing treatment patterns and results beyond high-volume academic centers, thereby reducing selection bias commonly encountered in single-center or registry-based studies.

Another strength lies in the inclusion of a wide range of contemporary treatment techniques—both established and emerging—allowing a nuanced comparison of EVT modalities (SC, BAC, SAC, FD, IFD) and NSC in ruptured and unruptured aneurysms. The use of propensity score weighting further improves comparability between treatment groups by adjusting for baseline differences such as age, sex, comorbidities, and treatment year.

However, limitations inherent to administrative data must be acknowledged. Most notably, the lack of detailed clinical and anatomical information (e.g., aneurysm size, location, morphology, or clinical grading scores) limits the ability to fully adjust for case complexity and treatment feasibility. Furthermore, only in-hospital outcomes are available, precluding the assessment of long-term efficacy, retreatment, or rebleeding rates. For making balanced treatment decisions, improvement of the initially lower functional outcomes at hospital discharge after NSC (following rehabilitation at 3-6 months) together with higher long-term aneurysm occlusion rates compared to SC need to be considered.

Coding accuracy may also vary across institutions, particularly for newer or less commonly used devices, which may affect the reliability of procedural categorization. Additionally, patients undergoing multiple procedures or inter-hospital transfers may be double-counted, leading to potential overestimation in specific groups.

Lastly, while EVT is often associated with lower short-term resource use, the higher procedural costs of advanced devices (e.g., FD, IFD) and the lack of long-term follow-up limit conclusions on overall cost-effectiveness. A comprehensive economic evaluation incorporating both initial and downstream costs remains necessary.

Despite these limitations, the study offers robust and contemporary insights into the in-hospital performance of evolving aneurysm treatment strategies across a national population.

## Conclusion

This nationwide analysis of in-hospital outcomes highlights significant differences among treatment modalities for IAs. SC remains the most frequently used and effective therapy for ruptured IAs, offering superior short-term functional outcomes and lower mortality compared to NSC, SAC, and FD. BAC and IFD demonstrated comparable safety to SC in this setting. For unruptured IAs, BAC, SAC, and especially IFD were associated with improved functional independence. NSC, by contrast, yielded less favorable short-term outcomes. Despite limited utilization, IFD showed the most favorable in-hospital profile—including better functional independence, shorter ventilation time, and reduced hospital stays.

While SC continues to be the standard of care, these findings support the safe application of IFDs for anatomically suitable aneurysms, particularly wide-neck bifurcation types. Future studies should focus on long-term outcomes, retreatment rates, and cost-effectiveness to better define the optimal therapeutic strategy for different aneurysm subtypes.

## Data Availability

Starting from 2005, the German Federal Statistical Office (DESTATIS) has been providing summarized and anonymous results concerning specific inquiries related to billing data in Germany's hospitals. This information is based on the diagnosis related group system and encompasses diagnoses (classified under the International Classification of Diseases, Tenth Revision, ICD-10) as well as performed procedures (coded according to the Operation and Procedure Classification System). These data are accessible upon request and can be utilized by anyone, if they adhere to the specified terms of use. To this study, only these publicly available and aggregated routine health data were examined.

## Acknowledgments

None

## Supplemental Material

Tables S1–S6

## Nonstandard abbreviations and acronyms

|  |  |
| --- | --- |
| BAC | balloon-assisted coiling |
| BRAT | Barrow ruptured aneurysm trial |
| DAPT | dual antiplatelet therapy |
| DESTATIS | Deutsches Statistik-Informationssystem (German federal statistical office) |
| DRG | diagnosis related group |
| EVT | endovascular treatment |
| FD | flow-diversion |
| GCP | good clinical practice |
| GDC | Guglielmi detachable coiling |
| IA | intracranial aneurysm |
| ICD-10 | International Classification of Diseases, 10. Revision |
| ICH | intracerebral hemorrhage |
| IFD | intrasaccular flow disruption |
| ISAT | international subarachnoid aneurysm trial |
| mRS | modified Rankin scale |
| NIS | nationwide inpatient sample |
| NIS-SOM | nationwide inpatient sample-subarachnoid hemorrhage outcome measure |
| NIS-SSS | nationwide inpatient sample subarachnoid hemorrhage severity score |
| NSC | neurosurgical clipping |
| OPS | operation and procedure classification system |
| SAPT | single antiplatelet therapy |
| SAC | stent-assisted coiling |
| SAH | subarachnoid hemorrhage |
| SC | standard coiling |
| SMD | standardized mean difference |

